# Effectiveness of community-based burden estimation to achieve elimination of lymphatic filariasis: a comparative cross-sectional investigation in Côte d’Ivoire

**DOI:** 10.1101/2022.02.10.22270792

**Authors:** Hope Simpson, Daniele O. Konan, Kouma Brahima, Jeanne d’Arc Koffi, Saidi Kashindi, Melissa Edmiston, Stefanie Weiland, Katherine Halliday, Rachel Pullan, Aboulaye Meite, Benjamin Guibehi Koudou, Joseph Timothy

## Abstract

For lymphatic filariasis (LF) elimination, endemic countries must document the burden of LF morbidity (LFM). Community-based screening (CBS) is used to collect morbidity data, but evidence demonstrating its reliability is limited. Recent pilots of CBS for LFM alongside mass drug administration (MDA) in Côte d’Ivoire suggested low LFM prevalence (2.1-2.2 per 10,000).

We estimated LFM prevalence in Bongouanou District, Côte d’Ivoire, using a comparative cross-sectional design. We compared CBS implemented independently of MDA, adapted from existing Ministry of Health protocols, to a population-based prevalence survey led by formally trained nurses. We evaluated the reliability of case identification, coverage, equity, and cost of CBS.

CBS identified 87.4 cases of LFM per 10,000; the survey identified 47.5 (39.4-56.3; prevalence ratio [PR] 1.84; 95% CI 1.64-2.07). CBS identified 39.7 cases of suspect lymphoedema per 10,000; the survey confirmed 35.1 (29.2-41.5) filarial lymphoedema cases per 10,000 (PR 1.13 [0.98-1.31]). CBS identified 100.3 scrotal swellings per 10,000; the survey found 61.5 (55.5-67.8; PR 1.63 [1.41-1.88]); including 26.6 (21.5-32.4) filarial hydrocoele per 10,000 (PR of suspect to confirmed hydrocele 3.77 [3.12-4.64]). Positive predictive values for case identification through CBS were 64.0% (54.5-72.8%) for filarial lymphoedema; 93.2% (88.5-96.4%) for scrotal swellings; and 33.3% (26.4-40.8%) for filarial hydrocoele. Households of lower socioeconomic status and certain minority languages were at risk of exclusion. Direct financial costs were $0.17 per individual targeted and $69.62 per case confirmed. We provide our CBS toolkit.

Our community-based approach to LFM burden estimation appears scalable and provided reliable prevalence estimates for LFM, scrotal swellings and LF-lymphoedema. The results represent a step-change improvement on CBS integrated with MDA, whilst remaining at programmatically feasible costs. Filarial hydrocoele cases were overestimated, attributable to the use of case definitions suitable for mass-screening by informal staff. Our findings are broadly applicable to countries aiming for LF elimination using CBS.

**Summary box:** *What is already known?:*

- In many lymphatic filariasis (LF) endemic countries, community-based screening (CBS) is used alongside mass drug administration (MDA) campaigns to estimate the burden of disease (LFM), required to achieve WHO targets for disease elimination.
- Previous studies have shown that the accuracy of CBS for LFM varies widely and the underlying contextual factors that impact on effectiveness remain unclear.
- In Côte d’Ivoire in 2020, pilot studies alongside MDA suggested low prevalence of LFM, though a process evaluation indicated estimates were negatively affected due to the competing demands of MDA, a challenge reported in other settings.

*What are the new findings:*

- We strengthened CBS in Bongouanou Department, Côte d’Ivoire, and de-coupled activities from MDA, which led to a 40-fold increase in LFM case estimates. We validated this estimate using a population-based prevalence survey led by formal healthcare workers, which demonstrated a comparable estimate of LFM.
- The direct financial cost of CBS was comparable to, or less than, other large-scale NTD interventions and supports scalability as a programmatic activity.
- We quantified specific biases of CBS, including poor differentiation between hydrocoele and scrotum swellings of alternative aetiology, and preferential inclusion of household based on sociodemographic characteristics.

*What do the new findings imply:*

- The approach we developed for strengthened, standalone CBS can provide estimates of LFM that reflect the true burden of disease, and is applicable to other LF endemic countries utilising large-scale community-based approaches.
- There is likely to be a high burden of LFM in endemic districts requiring expansion of morbidity management and disability prevention services.
- Accurate delineation of hydrocoele from other causes of scrotal swellings appears unfeasible using informal cadres employed during CBS. In settings where CBS is implemented, health providers should consider integration of conditions in activities.

*Public Involvement:* Two CDDs who had been involved in pilot LFM screening activities in 2020 participated in a review workshop, providing feedback which was used to develop the toolkit for LFM screening. Prior to the start of study activities, a launch meeting was held with traditional and religious leaders, administrative authorities and representatives of men’s, women’s and children’s groups of Bongouanou district. Participants were asked for their perspective on the importance of LFM as a health problem in their communities. Through this meeting they were informed about the CBS and nurse-led survey, and asked to cascade information through their communities. Patients were not involved in the study design.

## Introduction

Lymphatic filariasis (LF) is a mosquito-borne disease caused by the filarial nematodes *Wuchereria bancrofti, Brugia malayi* and *Brugia timori* [1], estimated to infect 51 million people globally [2]. Progression of the infection to chronic disease is associated with progressive damage to the lymphatic system, which can lead to irreversible swelling and acute attacks of dermato-lymphangio-adenitis (ADLA). The two most overt manifestations of chronic LF are lymphoedema-caused by accumulation of lymph fluid in the soft tissue, generally affecting a limb or breast- and hydrocoele, caused by accumulation of lymph fluid inside the scrotal sac [3]. Although the number of people affected by LFM is difficult to quantify (data being hard to collect and predictions unreliable), the burden of disease is undoubtedly substantial— estimates from 2012 suggested 19.4 million men with filarial hydrocoele and 16.7 million people with lymphoedema globally [4]. Disability-adjusted life year (DALY) estimates place LF as the second leading cause of disability due to parasitic diseases [5].

Since 1997, LF has been targeted for elimination as a public health problem, with efforts coordinated by WHO under the Global Programme to Eliminate Lymphatic Filariasis (GPELF) [6]. The elimination strategy consists of two components: interruption of transmission through mass drug administration (MDA) in endemic areas, and alleviation of suffering through morbidity management and disability prevention (MMDP) [7]. Verification of elimination depends on the sustained reduction of prevalence to below 1%, documentation of lymphoedema and hydrocoele case numbers, and the readiness and quality of MMDP services in health facilities in endemic areas [8]. While MDA has helped reduce LF infection prevalence, many people remain affected by lymphoedema and hydrocoele and require these services. However, most endemic countries lack estimates of the numbers of people affected and their distribution. There is no standardised mechanism for the collection of data on LFM, but due to resource limitations and barriers to care-seeking at health facilities [9-11], case detection in sub-Saharan Africa often depends upon existing community health infrastructure [12]. Community-based interventions, particularly MDA, have been central to the control of NTDs in sub-Saharan Africa (SSA), enabling access to marginalised communities and higher intervention coverage [13-15], and have been successfully used for case-finding in Guinea worm eradication [16]. The suitability of these staff to identify and refer cases of more subtle, chronic NTD morbidity is less certain, however. Community-based screening (CBS) of LFM during MDA is used in Burkina Faso, Ghana and Malawi [17] and other SSA countries, but is understood to substantially underestimate true case numbers [18].

In Côte d’Ivoire, where LF is endemic in 99 of 103 health districts and infection prevalence is predicted to exceed 1% [2], the NTD Control Programme (NTDP) plans to estimate LFM burden using CBS to guide service delivery and achieve LF elimination. The system will be scaled up gradually, providing opportunity to iteratively tailor and strengthen it. CBS through MDA piloted in 2020 identified 20 cases of lymphoedema and 9 of hydrocoele in the district of Lakota (population >=15 years old 132,216), and 34 cases of lymphoedema and 17 of hydrocoele in the district of Divo (population >=15 years old 248,613) [19,20]. We aimed to improve this existing approach by de-coupling case finding from MDA, improving and strengthening the CDD training programme, and implementing simple methodological adaptations. We evaluated the effectiveness of this strengthened, standalone strategy against a population-based prevalence survey led by nurses specially trained in LFM diagnosis. We also conducted a process evaluation during implementation to understand operational factors affecting performance including diagnostic reliability, household coverage, equity, and financial cost.

## Methods

### Setting

The study was conducted in the health district of Bongouanou, in the Moronou Region of centre-east Côte d’Ivoire (Figure 1S). The district is LF-endemic, with evidence from geospatial and models and a community-based survey using ICT cards in 2012 suggesting high prevalence of parasitaemia [2,21,22], but had not previously been surveyed for LFM. The landcover is primarily Guinean forest-savanna mosaic and the main industry is cacao and coffee farming [23]. Compared to other districts in the region, the infant mortality rate is low, public and private-sector salaries are high, and there are high levels of access to piped water [23]. The district population used for health operation purposes including MDA, is 169,999, the population >=15 years old is 94,319 and the male fraction is 0.50 [19]. The district is serviced by 24 health facilities: one general hospital, four urban health centres, eight rural health centres, and ten dispensaries. The area covered by each health facility is referred to as a health area, and populations within these health areas range from 800-26,513.

**Figure 1:**
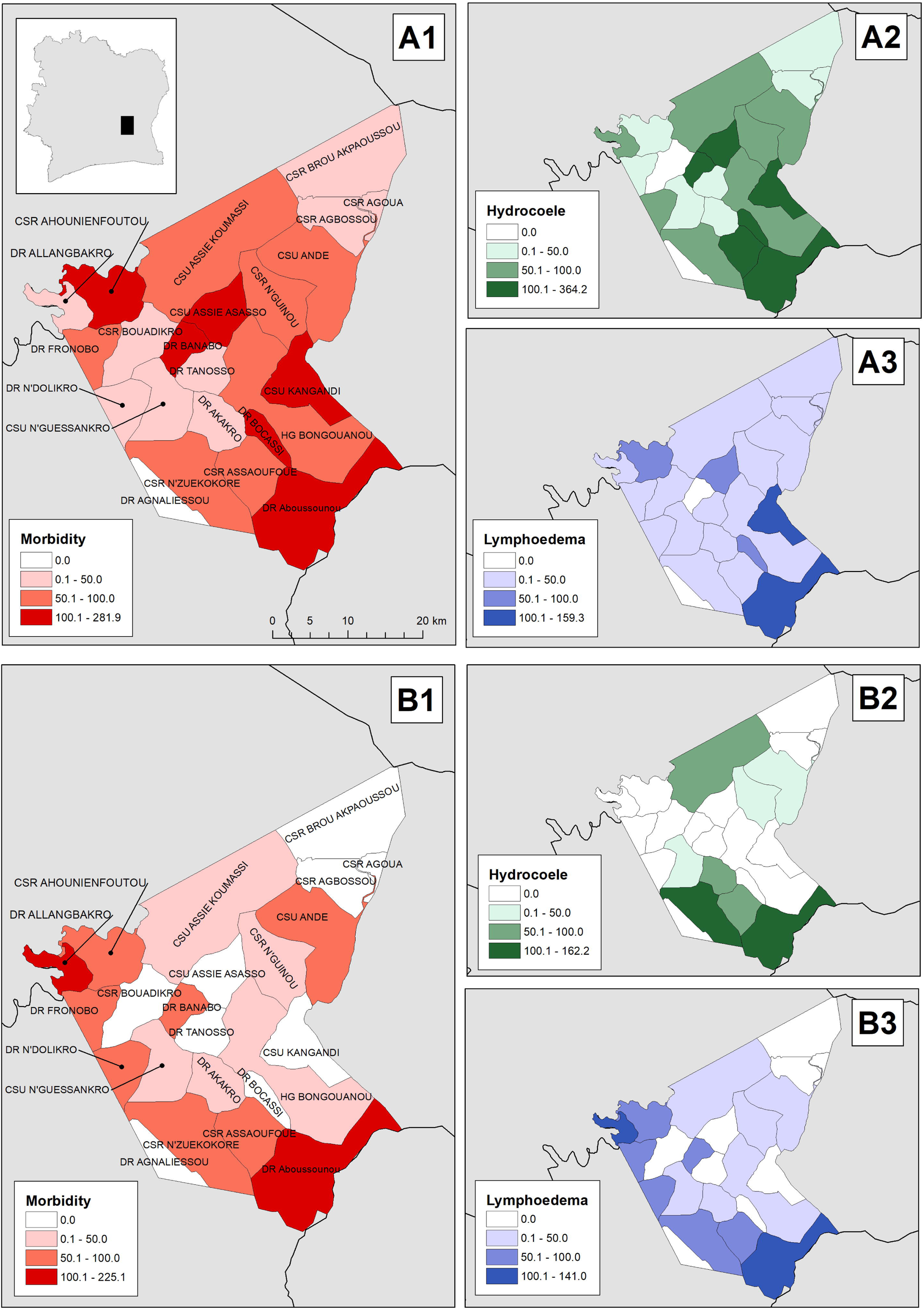
Prevalence (number of cases per 10,000 population) of 1) lymphatic filariasis morbidity, 2) filarial lymphoedema, and 3) hydrocoele, detected through A) community-based screening led by volunteers and B) population-based prevalence survey led by formally trained nurses in Bongouanou, Côte d’Ivoire.

### Study design and participants

We did a comparative cross-sectional study of methods to estimate LFM prevalence. Between 22^nd^ and 26^th^ February 2021, all LF-MDA community drug distributors (CDDs) completed a stand-alone, exhaustive door-to-door case search for leg swellings (suspect lymphoedema) and scrotal swellings (suspect hydrocoele) covering the entire population aged 15 years and over. Results were compared to estimates from a population-based prevalence survey, led by nurses specially trained in diagnosis of LFM, conducted immediately after (15^th^ March-17^th^ June 2021).

For the survey, we used a stratified two-stage cluster-based design with strata based upon health areas, primary sampling units (PSUs) defined as CDD zones and secondary sampling units (SSUs) as households [24]. Total PSUs selected within strata was determined using proportional allocation. PSUs were selected using simple random sampling without replacement, due to absence of population data at CDD zone level. All individuals aged 15 years and older in selected PSUs were eligible for participation. Using a standard sample size calculation [24] assuming a prevalence of 5 cases per 1,000 population, a participation rate of 95%, design effect of 5.95, and applying a finite population correction factor for the population of Bongouanou district, we calculated that 12,217 participants needed to be examined to estimate LFM prevalence with an absolute precision of 0.003.

To assess diagnostic reliability of CBS, CDDs recruited all identified cases for re-examination by nurses at a central location after completion of surveys within PSUs.

### Procedures

#### Co-development of toolkit for morbidity enumeration

The CDD toolkit, including the training of trainers guide, slide-deck, job-aid and photobook, was developed by the project team before being presented to a representative of the NTDP (BK) for further revisions. The draft materials were then extensively reviewed and modified at a 3-day revisions workshop held in Yamoussoukro, attended by the NTDP, members of the project team, and the team who had been involved in pilot CBS implemented by the MoH NTD programme in 2020 (members of the district health team, one CDD, and partners). The revised toolkit is available in the Supplementary File (study toolkit: community screening).

#### CDD training and community-based screening

Based on feedback from the pilot, we adapted existing approaches to enumeration. The main changes were that CBS was implemented as a standalone activity, separate from LF MDA, and that CDDs were instructed to exclusively use a door-to-door strategy and avoid fixed-post activities. A full list of modifications on the existing approach is shown in Table 1.

**Table 1:**
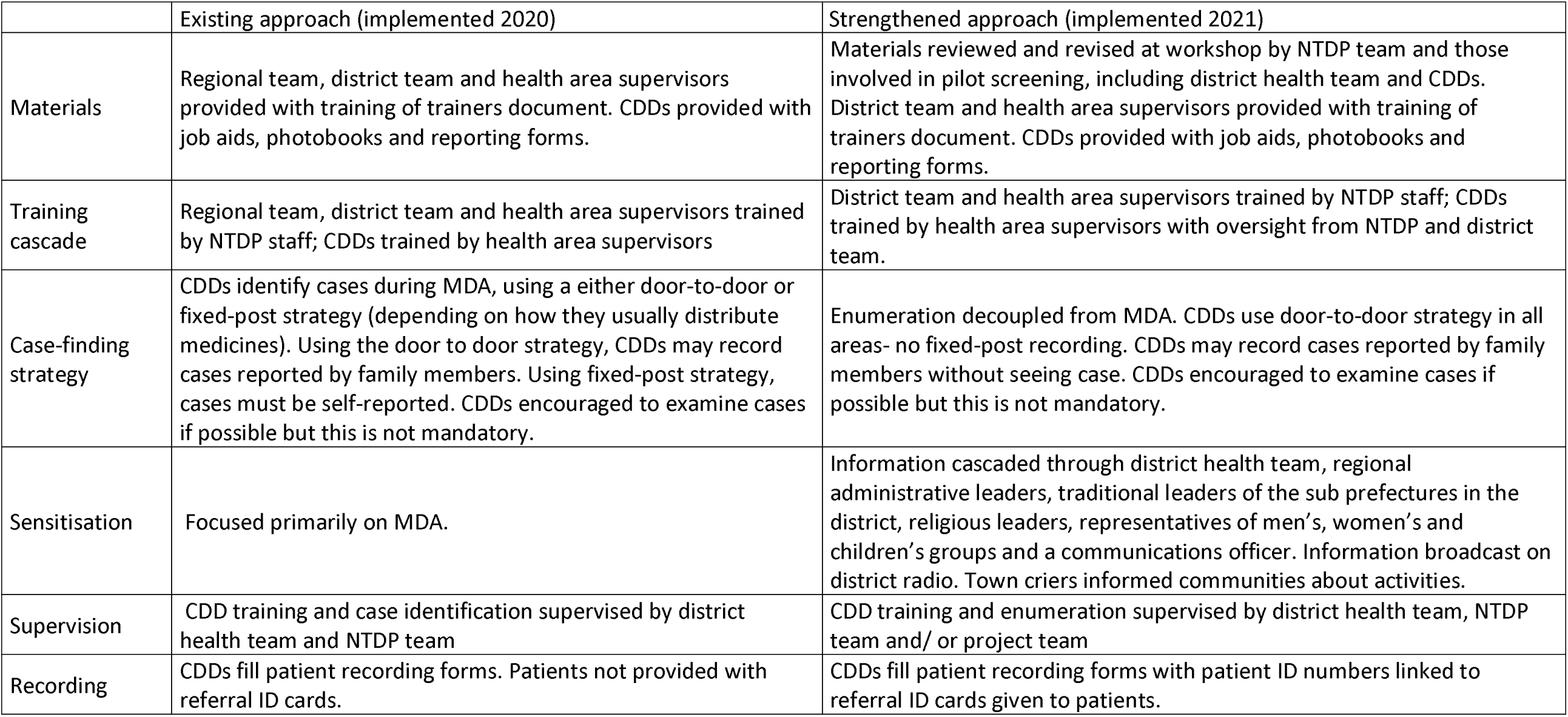
Existing and strengthened approach to community-based enumeration of lymphatic filariasis morbidity (LFM) in Côte d’Ivoire.

Training of CDDs was delivered through a cascade. In the first stage, health district staff and health area supervisors were trained by the head of MMDP in the NTDP (BK) using the slide deck and patient demonstrations. Health area supervisors then cascaded training to CDDs in their respective areas using the training of trainers guide, photobook and job aid, with practical demonstrations. Following training, CDDs undertook a post-training quiz which was evaluated by the project team in order to assess the quality of the training provided.

All identified cases were provided with unique patient ID cards for re-capture during evaluation surveys to enable case validation. CDDs recorded cases on paper forms including patient demographic and contact details and basic clinical information and medical history. Data from these forms was entered by trained supervisors into an electronic database via electronic devices running an ODK-based application.

### Nurse-led evaluation survey

We recruited health supervisors with nurse or midwife qualifications from Bongouanou district as clinical field surveyors. Twenty-six supervisors underwent a 3-day training programme on the diagnosis and management of lymphoedema and hydrocoele led by specialist and experienced dermatologists from the University Hospital of Treichville in Abidjan and by the head of the MMDP unit from the NTDP. Training materials are available in the Supplementary File (study toolkit: LFM evaluation survey). Participants were assessed through a post-training test, and 18 nurses were selected for implementation.

For household (SSU) selection in PSUs, teams of 3 nurses followed separate random walks beginning from randomised start points assigned by a custom built ODK-based application. An initial household census and interview was completed to collect information on sociodemographic variables, GPS location and CDD coverage using electronic devices running an ODK-based application. Each consenting individual was checked for swelling on the limbs, and males underwent a brief testicular examination. Suspect cases were defined according to the same case definitions used by CDDs, and underwent detailed examination for confirmatory diagnosis. If any eligible participants were absent, remaining eligible household members were shown pictures of LFM from a flipbook and acted as proxy respondents. Anyone identified through this screen was defined as a suspect case and targeted for follow-up examination.

### Outcomes

The CDD case definition for suspect lymphoedema was *an increase in the volume of a limb or breast in a person aged 15 years or older* and that for suspect hydrocoele was *swollen testicles in a male aged 15 years or older*.

The case definition of filarial lymphoedema was *swelling of limb or breast, in a patient aged 15 years older, present for at least a year but not since birth, and not due to leprosy, erysipelas, malignancy, surgery, or heart disease*. The definition of filarial hydrocoele was *a discrete, nontender mass around the testes, not explained by an inguinal hernia or scrotal lymphoedema, not present since birth and present for more than 24 hours*. Lower limb lymphoedema was classified according to the Dreyer system [25]. Cases of testicular swelling were characterised according to the system proposed by Capuano and Capuano [26].

Clinically confirmed cases of filarial lymphoedema and hydrocoele were given advice on self-care, a patient identification card, and re-imbursement for travel costs to the local health facility. Confirmed hydrocoele cases were registered for inclusion in planned hydrocoele surgery within the district.

### Statistics and data analysis

#### Prevalence estimation

We calculated the crude prevalence of suspect LFM detected by CDDs at district and health area levels using estimates of the district total and male population aged 15 years and older from the Côte d’Ivoire National Institute of Statistics as denominators [19]. Since PSU populations were not available from district health databases, design weights were assigned using population estimates extracted from the Facebook population density layer [13]. Further details are given in Supplementary file S1.1. To enable spatial delineation of CDD zones, nurses walked the boundary of PSUs with CDDs, capturing the geographical limits of the catchment using GPS-enabled devices [27]. We calculated district-level prevalence estimates of LFM outcomes using ***survey*** package in R (version 4.1-1) [28] with post-stratification weighting applied for age and sex [19]. Prevalence estimates of LFM outcomes from CDD enumeration were compared to household survey prevalence estimates using risk ratios.

To understand contextual and operational factors affecting CDD coverage, we developed a mixed-effects generalized linear model (binomial distribution), for reported visitation by CDDs. We assessed sociodemographic variables at household level, including a multi-dimensional indicator of socioeconomic status (SES) constructed using latent class analysis (LCA; full details in the Supplementary File S1.2) and PSU-level indicators of CDD demographics, performance and community accessibility (All candidate predictors and sources are shown in Table 1S). Continuous variables were centred and scaled. Missing data were imputed by single imputation of PSU means or modes (for continuous and categorical variables respectively). Random intercepts were allowed for CDD zones nested within health areas. Candidate variables were subject to bivariate analysis and included within final models if p <=0.2 using likelihood ratio tests, given large parameter space and absence of observed collinearity [29]. Final models were assessed for violations of assumptions.

To estimate positive predictive value (PPV; the proportion of identified cases confirmed by gold standard diagnosis [30]) of case identification by CDDs, CDD-identified LFM cases were linked to patients examined by nurses using capture re-capture of coded patient ID cards. We estimated PPV using ***epiR*** [31], with confirmed diagnosis by a trained nurse as gold-standard.

We estimated the direct financial costs of CDD case finding using an ingredients-based approach to estimate costs per person targeted by the screening activity and per confirmed case identified. Costs were categorised by phase of activity (sensitisation, training of trainers (first stage of cascade), training of CDDs (second stage of cascade) and enumeration.

### Ethics

Eligible participants (those aged 15 years and older) were provided with an information sheet and the study was explained. Written informed consent was obtained from individuals aged 18 years and above. Minors (aged <18 years) provided oral assent, and written informed consent was obtained from their parents or legal guardians. The study was granted ethical approval by Le Comité National d’Ethique des Sciences de la Vie et de la Santé in Côte d’Ivoire and the Ethics Committee of the London School of Hygiene and Tropical Medicine (Reference 21203).

## Results

### Study participants

Across 110 PSUs, nurses visited 8,247 households which were occupied and had an adult present. Of these, 58 (0.70%) refused to participate. Across 8,189 households, 12,289 people were invited to participate and 12,287 (99.99%) agreed, including 4,818 males (39.2%). Twenty participants (0.16%) refused limb examinations and 202 males (4.2%) refused scrotal examination. The median number of households visited per cluster was 75 (interquartile range [IQR] 61-86), and the median number examined per cluster was 98 (IQR 77-112).

### Reliability of community-based screening

Following an exhaustive case search across the district among people >=15 years old (population 94,319; 47,349 males), CDDs identified 824 suspect LFM cases: 374 lymphoedema, 475 hydrocoele, and 25 with both (Table 2). The prevalence of suspect LFM was 87.4 per 10,000 (95% CI 81.5-93.5 per 10,000). The prevalence of suspect lymphoedema was 39.7 per 10,000 (95% CI 35.7-43.9 per 10,000) and that of suspect hydrocoele 100.3 per 10,000 males (95% CI 91.5-109.7 per 10,000) (Figure 1).

**Table 2:**
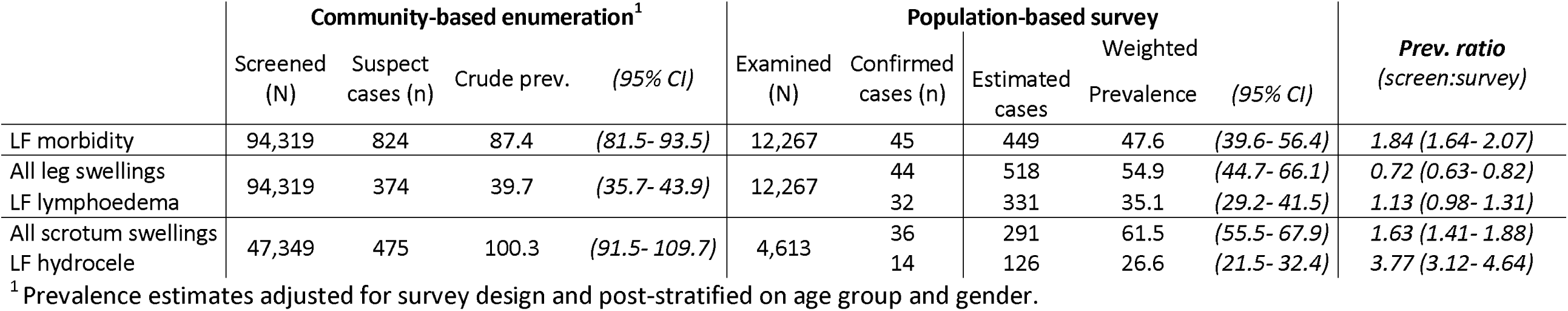
Number and prevalence (per 10,000) of LFM cases detected through community-based enumeration and population-based survey.

In the population-based survey, 45 cases of LFM were confirmed, giving a prevalence of 47.6 per 10,000 (95% CI 39.6-56.4) after adjusting for survey design and non-response. The prevalence ratio of CDD-identified suspect LFM to confirmed LFM was 1.84 (95% CI 1.64-2.07). Forty-four cases of leg swellings were identified and 32 were confirmed as filarial lymphoedema. The design-adjusted prevalence estimate of confirmed filarial lymphoedema was 35.1 (95% CI 29.2-41.5) per 10,000; within confidence bounds of the CDD estimate (prevalence ratio of 1.13 [95% CI 0.98-1.31]). Thirty-six cases of scrotal swellings were identified and 14 confirmed as filarial hydrocoele. The adjusted prevalence of scrotal swellings was 61.5 per 10,000 males (95% CI 55.5-67.9). The prevalence ratio for scrotal swellings was 1.63 (95% CI 1.41-1.88). The prevalence of confirmed filarial hydrocoele was 26.6 per 10,000 (95% CI 21.5-32.4), with a prevalence ratio of 3.77 (95% CI 3.12-4.64). One patient presented both filarial lymphoedema and hydrocoele.

### Cases missed by CDDs

Of 32 confirmed filarial lymphoedema cases identified in the nurse-led household survey, 10 (31.3%) had not been identified despite the household having been visited. Of 36 cases of scrotal swellings identified in the household survey, 6 (16.7%) had been missed despite their household being visited. The low number of missed cases limited precise comparisons between those missed and identified.

### Reliability of LFM case identification by CDDs

To assess reliability of case identification, nurses examined cases identified by CDDs at post-survey clinics. Of 374 suspect lymphoedema cases, 114 (30.5%) were re-examined and leg swellings were confirmed in 88 cases, giving a PPV of 77.2% (96% CI 68.4-84.5%). Filarial lymphoedema was confirmed in 73 of the 114 cases (PPV 64.0%, 95% CI 54.5-72.8%). The 15 cases with leg swelling not diagnosed as filarial lymphoedema included 4 cases of Buruli ulcer, 3 of erysipelas, and 9 other diagnoses.

Of 475 suspect hydrocoele cases identified by CDDs, 177 (37.3%) underwent scrotal examination by nurses. Scrotal swelling was confirmed in 165. The PPV for identification of scrotal swellings was 93.2% (95% CI 88.5-96.4%). Filarial hydrocoele was confirmed in 59 cases (PPV 33.3%, 95% CI 26.4-40.8%). Among the 106 non-filarial scrotal swellings, 97 were cases of hernia and 9 were other diagnoses. It is important to note that CDDs were not asked to differentiate between filarial and non-filarial aetiology for suspected case definitions (see Methods).

### Household coverage and equity of community-based screening

Of 8,189 households interviewed by nurses, 5,265 (64.3%; 95% CI 63.2-65.3%) reported being visited by a CDD during CBS; 2,769 (33.8%) had not; and 154 (1.9%) did not know. Using causal modelling approaches to account for household, PSU and health-area contextual factors, household size was positively associated with inclusion in the CBS (Table 3). Households in which the primary language of the household head was Baoule, Senoufo, Dioula or other languages were less likely to have been visited than those in the predominant ethno-lingual group of Bongouanou, Agni. Households owning a basic mobile or smartphone were also more likely to have been visited than those without, independent of socioeconomic status. Crudely, households in the lowest socioeconomic class based on LCA were more likely to have been visited than those in the middle and highest classes, but after adjusting for survey design and other covariates, the middle SES class was more likely to have been visited. PSU-level fixed effects suggested higher coverage in more socioeconomically-developed clusters, with a positive association of household coverage with stable night light. A very low proportion of variance was attributable to health area levels (ICC L1 = 0.099) indicating little effect of implementation at this scale.

**Table 3:** Predictors of household inclusion in community case search.

| Predictors of visitation by CDDs |  | n visited by CDD | N surveyed by nurses | Risk (%) | Multivariate Analysis |  |
| --- | --- | --- | --- | --- | --- | --- |
| Household-level Fixed Effects |  |  |  |  | Adjusted OR (95% CI) | LRT p-value |
| Log household size |  |  |  |  | 1.07 (1.06- 1.09) | <0.001 |
| Phone ownership<sup>1</sup> |  |  |  |  |  | <0.001 |
|  | No phone | 747 | 1,303 | 57.33 | 1 (base) |  |
|  | Basic phone only | 740 | 1,156 | 64.01 | 1.11 (1.07- 1.15) |  |
|  | Smartphone | 3,645 | 5,419 | 67.26 | 1.06 (1.03- 1.09) |  |
| Language<sup>2</sup> |  |  |  |  |  | <0.001 |
|  | Agni | 3,998 | 6,022 | 66.39 | 1 (base) |  |
|  | Baoule | 322 | 511 | 63.01 | 0.94 (0.91- 0.98) |  |
|  | Dan | 10 | 21 | 47.62 | 0.94 (0.78- 1.12) |  |
|  | Dioula | 287 | 492 | 58.33 | 0.93 (0.90- 0.97) |  |
|  | French | 18 | 33 | 54.55 | 1.18 (1.01- 1.38) |  |
|  | Malinke | 110 | 160 | 68.75 | 0.96 (0.89- 1.02) |  |
|  | Senoufo | 33 | 59 | 55.93 | 0.88 (0.79- 0.98) |  |
|  | Other/ Unknown | 354 | 580 | 61.03 | 0.92 (0.89- 0.96) |  |
| Socioeconomic status<sup>3</sup> |  |  |  |  |  | 0.001 |
|  | Lowest | 504 | 658 | 76.6 | 1 (base) |  |
|  | Middle | 2,925 | 4,284 | 68.28 | 1.06 (1.01- 1.10) |  |
|  | Highest | 1,703 | 2,936 | 58 | 1.02 (0.98- 1.07) |  |
| Cluster/CDD-level Fixed Effects |  |  |  |  |  |  |
| CDD score<sup>4</sup> |  |  |  |  | 1.04 (1.00- 1.08) | 0.058 |
| CDD gender |  |  |  |  |  | 0.083 |
|  | Female | 1,722 | 2,474 | 69.6 | 1 (base) |  |
|  | Male | 3,410 | 5,404 | 63.1 | 0.93 (0.86- 1.01) |  |
| Mean distance to stable lights |  |  |  |  | 1.06 (1.00- 1.12) | 0.047 |
| Mean accessibility to HFs |  |  |  |  | 1.03 (0.99- 1.07) | 0.123 |
|  |  |  |  |  | ICC |  |
|  |  |  |  |  | CDD zones within health areas (L1:L2) | 0.099 |
|  |  |  |  |  | CDD zones (L2) | 0.145 |

<sup>1</sup> Missing for 1 household. <sup>2</sup> Missing for 9 households. <sup>3</sup> Multidimensional indicator constructed from: household electricity connection (missing for 5 households), education of household head (missing for 1 household), dwelling walls made from improved material (missing for 3 households), dwelling floor made from improved material (missing for 1 household), household access to improved water supply (missing for 3 households). Households missing data were assigned the cluster modal value. <sup>4</sup> Missing for 3 CDDs equating to 156 households, which were not included in this model.

**Table 4:** Demographic and clinical characteristics of confirmed cases of lymphatic filariasis morbidity identified.

| Demographic/ clinical characteristics | Confirmed filarial lymphoedema (n=121) |  |  | Demographic/ clinical characteristics | Confirmed filarial hydrocoele (n=107) |  |  |
| --- | --- | --- | --- | --- | --- | --- | --- |
|  | N | % / median | CI/IQR |  | N | % / median | CI/IQR |
| **Gender** |  |  |  |  |  |  |  |
| Female | 71 | 58.7 | 49.4- 67.6% |  |  |  |  |
| Male | 50 | 41.3 | 32.4- 50.6 % |  |  |  |  |
| **Age (median)** |  | 47 | 34.0- 57.0 | **Age (median)** |  | 51 | 40.0- 64.0 |
| **Years of swelling (median)** |  | 6.5 | 3.0-15.0 | **Years of swelling (median)** |  | 5 | 3.0-10.8 |
| **Location of swelling** |  |  |  |  |  |  |  |
| Upper limb, unilateral | 4 | 3.3 | 0.9- 8.2% |  |  |  |  |
| Lower limb, unilateral | 73 | 60.3 | 51.0- 69.1% |  |  |  |  |
| Lower limb, bilateral | 42 | 34.7 | 26.3- 43.9% |  |  |  |  |
| One upper & one lower limb | 2 | 1.7 | 0.2- 5.8% |  |  |  |  |
| **ADLA Frequency** |  |  |  | **ADLA Frequency** |  |  |  |
| Never | 40 | 33.1 | 24.8- 42.2% | Never | 67 | 55.4 | 52.7- 71.8% |
| Less than once per month | 54 | 44.6 | 35.6- 53.9% | Less than | 26 | 21.5 | 15.5- 33.5% |
| At least once per month | 25 | 20.7 | 13.8- 29.0% | At least once | 13 | 10.7 | 6.6- 19.9% |
| Unknown | 2 | 1.7 | 0.2- 5.8% | Unknown | 1 | 0.8 | 0.02- 5.1% |
| **Dreyer Stage<sup>1</sup> (lower limb only)** |  |  |  | **Stage<sup>2</sup>** |  |  |  |
| 1- 2 | 78 | 66.7 | 55.2- 73.0% | 1 | 21 | 17.9 | 12.6- 28.4% |
| 3+ | 39 | 33.3 | 24.0- 41.3% | 2+ | 84 | 71.8 | 69.5- 85.9% |
|  |  |  |  | Unknown | 2 | 1.7 | 0.2- 6.6% |
| **Entry lesions** |  |  |  | **Grade<sup>2</sup>** |  |  |  |
| Present | 38 | 31.4 | 23.3- 40.5 | 0 | 93 | 76.9 | 79.0- 92.7% |
| None | 83 | 68.6 | 59.5- 76.2% | 1+ | 11 | 9.1 | 5.2- 17.7% |
|  |  |  |  | Unknown | 3 | 2.6 | 0.6- 8.0% |

<sup>1</sup> According to the Dreyer system [25]: stage 1 = reversible lymphoedema (limb may return to normal, for example at night), stage 2+ = non-reversible lymphoedema.
<sup>2</sup> According to the staging and grading system of Capuano and Capuano [26]: Stage 1 = smaller than a tennis ball. Grade 0 = No visible burial or shortening of penis

### Patient Clinical Characteristics

Of the 121 confirmed cases of filarial lymphoedema, the majority (58.7%) were female and the median age was 47.0 years (interquartile range; IQR 34.0-57.0) (Table 2S). Most (60.3%) had unilateral lower limb swelling and the median duration of swelling was 6.5 years. Approximately a third (33.1%) had never experienced an acute attack, while 44.6% experienced less than one per month and 20.7 experienced at least one per month. Two thirds (66.7%) of cases were Dreyer stage one or two.

The median age of the 107 confirmed filarial hydrocoele cases was 51.0 years (IQR 40.0-64.0), and the median duration of swelling was 5.0 years (IQR 3.0-10.8). Most cases (55.4%) said they never experienced acute attacks, 21.5% experienced at least one per month and 10.7% experienced at least one per month. By the staging and grading system proposed by Capuano and Capuano [26], most cases (71.8%) were classified beyond stage 2 (scrotum larger than a tennis ball), but few (9.1%) were beyond grade 1 (visible burial of the penis).

### Financial Costs

The overall direct financial cost of CBS was 26,678.36 USD, of which approximately half was spent on preparation (sensitisation, training of trainers and of CDDs) and half on door-to-door screening by CDDs (Figure 2S). The cost per suspect case identified by CDDs was $33.60, that per case confirmed was $69.62 and that per person targeted was $0.17.

## Discussion

In this study, we evaluated a community-based strategy to estimate LFM burden, which can be implemented as a scalable, programmatic activity to support elimination of LF as a public health problem [8]. The strategy was programmatically feasible in terms of cost per person examined, and provided comparable prevalence estimates relative to a rigorous population-based survey. These were 40 times higher than those obtained in recent pilots implemented during MDA in Côte d’Ivoire. Taken together, these results indicate that the strengthened, standalone strategy appears a considerably more effective approach to describe the true epidemiological situation of LFM. We also quantified the coverage and equity of our community-based strategy, and identified challenges faced by CDDs in differentiating hydrocoele from scrotal swellings of other causes. Whether this latter challenge can be overcome cost-effectively remains an open question for the GPELF and LF-endemic countries.

We made a series of simple changes to the existing programmatic strategy that was piloted during MDA in 2020 in Côte d’Ivoire. The modifications included strengthening of the training programme, increased supervision, and de-coupling of case-finding from MDA. These appear to have facilitated a step-change improvement in LFM case enumeration, resulting in prevalence estimates 40 times higher than those from initial pilots [20]. All modifications are likely to have contributed to improved reliability of case detection, though further research, including process evaluation at scale, would be needed to elucidate the contribution of each element to success at different levels of implementation. We believe the de-coupling of case-finding from MDA was a significant enabler. Previous studies have shown dedicated case searches to be more effective than those embedded in other activities, suggesting that competing demands, rather than de-centralisation ***per-se***, are a more important barrier to reliable community-led LFM estimation [17,18].

Although the estimate of LFM prevalence detected by CDDs was higher than that shown by the prevalence survey, we consider the CDD-estimate reflective of the epidemiological situation in Bongouanou. CDDs appeared to face different challenges in the quantitation of suspect lymphedema and suspect hydrocele. Their identification of lymphoedema was good, though imperfect, in terms of both reliability and sensitivity: the PPV of 64% indicates that around one third of suspect cases identified by CDDs were not due to filarial lymphoedema, while the survey suggested that CDDs missed around a third of true cases (10 of 32 confirmed filarial lymphoedema cases had not been identified in the CBS). In effect, the similar magnitudes of these parameters resulted in an estimate of suspect LF-lymphoedema prevalence close to (within the confidence range of) that found in the evaluation survey. This is an encouraging result in terms of obtaining accurate estimates of lymphoedema prevalence through CBS, though the sensitivity of case identification could be further improved through modifications to the CDD training materials or community education to raise awareness of early signs of lymphoedema.

In contrast, the overestimate of scrotal swelling prevalence by CDDs (by around 63%) cannot be explained in terms of the PPV of case identification, which was 93.2% (indicating that fewer than 7% of suspect cases were misdiagnosed). One possible explanation for this discrepancy is that some cases recorded by CDDs were not true cases, but that those who were were more likely to present for follow-up examination, resulting in a biased estimate of PPV. Another possibility is that the CBS was more sensitive to scrotal swellings than the evaluation survey, since CDDs could record cases without physical examination while nurses could not. Among the 202 men who did not undergo scrotal examination during the household survey, there may have been cases who were willing to describe symptoms to a CDD but not to be examined by a nurse in the household setting, and thus went undetected.

The low PPV of hydrocoele identification led to an overestimation of hydrocoele prevalence by CDDs. This was not unanticipated, since CDDs were not trained to distinguish the aetiology of scrotal swellings, which was deemed unfeasible, particularly given the scale of coverage expected of them. A similarly low PPV of hydrocoele identification by CDDs has been demonstrated in Ghana, though the same study demonstrated a much higher PPV (92%) in Malawi [17]. Differentiation of hernia from hydrocoele may be possible with training [17], but given resource limitations within the GPELF, this may be unrealistic at scale. Under current funding structures, cases are only eligible for free surgical intervention if confirmed as filarial hydrocoele, despite the fact that the repair of groin hernia, by far the most prevalent alternative diagnosis in our study, is also resolved by a simple and cost-effective surgery [32]. Our findings reinforce the clear public health and economic arguments for integration of case finding and surgery for these conditions, which should receive consideration within LF endemic countries [33]. The reliability of lymphoedema identification by CDDs was much higher, with PPV similar to estimates from other settings [17].

An important consideration for our findings is the potential scalability of the approach. This is crucial as WHO elimination dossiers for LF necessitate exhaustive enumeration of LFM across all endemic and previously endemic areas, often covering very large populations. We believe the financial cost per person screened in our study was low from an NTD programmatic perspective, being comparable to the cost per person treated through MDA in African settings in the early 2000’s [34], and substantially lower than the cost per person examined in the global trachoma mapping project ($4.20 in Côte d’Ivoire in 2015) [35]. The cost per case confirmed was lower than published estimates of the cost per case found in community-based leprosy screening, varying from $72 (in Mali, 1999)-$313 (in Nigeria, 2002) [36]. The costs were also low relative to the financial burden on patients unable to work due to LFM, estimated at around $700 per individual affected [37], and costing almost $1.3 billion a year in lost productivity globally [38]. Hydrocoele surgeries are extremely cost-effective [39], and lymphoedema management costs far less than the economic benefits to patients over their lifetime [40]. Taken together, these results present a strong case for investment in community-led approaches to identify LFM cases to be linked to MMDP services. Parallel investment in universal health coverage and surveillance strengthening will be required for the sustainability of these programmes.

The level of household coverage achieved by CDDs was high, aligning to WHO coverage targets for LF MDA [41]. However, we identified factors at household and community (CDD zone) level that affected the probability of household inclusion. Within communities, households in the middle socioeconomic class were more likely to have been visited, while some minority language groups appeared at risk of exclusion. This may reflect CDD bias towards friends or those of specific social standing, which has been demonstrated in the context of MDA in Uganda [42]. To address this, the inclusion and sensitisation of minority language groups should be planned from early stages, and communications materials may need to be translated into multiple languages before implementation. There was a low level of variability at health area level, which supports the quality of the training cascade, given that CDDs were trained through supervisors at health area level.

Our study had several limitations. Both case-finding methods were fallible to ascertainment bias, as males, students, and people of working age would be less likely to be at home. Although we adjusted the survey prevalence estimate to correct this bias, it was not possible to do the same for the community-based estimate. Further, people affected by LFM may have been more likely to be at home-either because of unemployment or because they stayed intentionally for the survey. Whilst our evaluation survey was conducted following specialist clinical training, outcome measures were based on clinical diagnosis made by non-physician healthcare workers, which may be imperfect. Selection bias may have been introduced by random walk procedures. While we aimed to nullify this by using multiple, random start points, random walk is more prone to selection bias than fully randomised or segment-based sampling [43].

We have demonstrated the effectiveness of a scalable, community-based case-finding approach to LFM burden estimation. Together with our findings, the extensive toolkit we present can support programmes planning to implement similar activities. We believe this study transparently demonstrates how community-based infrastructure can support LF elimination, and is generalisable to other LF-endemic settings. Important advocacy points raised by our findings include the potential benefits of de-coupling LFM case-finding from MDA and the overt operational and public health benefits of integrating the detection and management of hydrocoele with scrotal hernia.

## Data Availability

Anonymised data will be made public upon publication of the manuscript

## Acknowledgements

We would like to thank the Bongouanou District Health Director and team for their support throughout the implementation of the study. We gratefully acknowledge the work of the CDDs and nurses who undertook the survey. We would also like to thank all patients and communities who participated in the survey. We would like to thank Dr. Steve Walker, LSHTM for assisting with the design of clinical training materials, and Prof Diabate and Dr Kouadio of the University of Treichville, Abidjan, for delivering the clinical training of field workers. Elliott Rogers contributed to the study toolkit, including the design of training materials and data collection forms.

